# Lockdown related travel behavior undermines the containment of SARS-CoV-2

**DOI:** 10.1101/2020.10.22.20217752

**Authors:** Nishant Kishore, Rebecca Kahn, Pamela P. Martinez, Pablo M. De Salazar, Ayesha S. Mahmud, Caroline O. Buckee

## Abstract

In response to the SARS-CoV-2 pandemic, unprecedented policies of travel restrictions and stay-at-home orders were enacted around the world. Ultimately, the public’s response to announcements of lockdowns - defined here as restrictions on both local movement or long distance travel - will determine how effective these kinds of interventions are. Here, we measure the impact of the announcement and implementation of lockdowns on human mobility patterns by analyzing aggregated mobility data from mobile phones. We find that following the announcement of lockdowns, both local and long distance movement increased. To examine how these behavioral responses to lockdown policies may contribute to epidemic spread, we developed a simple agent-based spatial model. We find that travel surges following announcements of lockdowns can increase seeding of the epidemic in rural areas, undermining the goal of the lockdown of preventing disease spread. Appropriate messaging surrounding the announcement of lockdowns and measures to decrease unnecessary travel are important for preventing these unintended consequences of lockdowns.

## Introduction

In response to the SARS-CoV-2 pandemic, unprecedented policies of travel restrictions and stay-at-home orders were enacted around the world almost simultaneously. These ranged from restrictions on human movement on a local scale to travel restrictions on regional and international scales. These policies were designed to reduce the spread of the SARS-CoV-2 virus by restricting the contact between infectious and susceptible individuals and to slow the spread of the virus out of epidemic hotspots.

Ultimately, the public’s response to announcements of lockdowns - defined here as restrictions on both local movement or long distance travel - will determine how effective these kinds of interventions are. Governments must give some warning to the public about upcoming travel restrictions to allow for necessary preparations, but a surge of travel prior to the lockdown being put in place risks the exact opposite of the desired effect, sending potentially infectious individuals out into previously unaffected regions around the country or internationally. In order to design effective policies in response to resurgence of SARS-CoV-2 or indeed in the context of future pandemics, understanding the human response to interventions is critical.

Analyses of aggregated data from mobile phones have been used to monitor movement patterns in the context of outbreaks [1, 2, 3], including this pandemic [4, 5]. Studies have shown that mobility patterns on local scales correlated with transmission within the city of Wuhan on a more local level [6, 7], and recent analyses have found associations between mobility and SARS-CoV-2 transmission in the United States [8]. Seasonal travel related to holidays, which creates a surge of travel out of cities, for example, can also have an important impact on the spread of infection [9]. Travel related to the Lunar New Year may well have spread the disease across China as the epidemic started to emerge in Wuhan. Since SARS-CoV-2 infected individuals are likely to be infectious prior to symptoms, and many may have no symptoms at all, the possibility of infected travelers unwittingly spreading the virus during large travel movements is significant.

Here, we measure the impact of the announcement and implementation of lockdowns on human mobility patterns by analyzing aggregated mobility data from mobile phones from multiple countries including India, France, Spain, and the USA, on local and national spatial scales. We show that immediately preceding lockdowns there was a consistent surge in travel out of the urban epicenters of the epidemic, or around local neighborhoods, in anticipation of restrictions. We observed urban-to-rural migration in each country analyzed, and we use a simple agent-based spatial model to examine how different behavioral responses to lockdown policies may spread epidemics, highlighting the importance of social considerations in the implementation of travel restrictions. We find that travel surges following announcements of lockdowns can increase seeding of the epidemic in rural areas, undermining the goal of the lockdown of preventing disease spread.

## Results

Subject to the availability of data (see Methods), we analyzed mobility patterns in response to lockdown orders on two spatial scales: i) within cities, and ii) more broadly on a national level.

### Pre-lockdown mobility surges and depopulation of cities

Lockdown announcements are generally made a few days before going into effect. To understand the impact of the announcement itself, we analyzed the percent change in population during the day compared to baseline in different parts of New York City based on Facebook data. As shown in Figure 1, Manhattan showed a large increase in population following the announcement during daytime hours, suggestive of mobility related to preparations for the lockdown, followed by a dramatic decline upon its implementation. In contrast, the population in most boroughs stayed the same or increased following the announcement and lockdown, consistent with stay-at-home orders that would have prevented people from commuting to other parts of the city for work.

**Figure 1.**
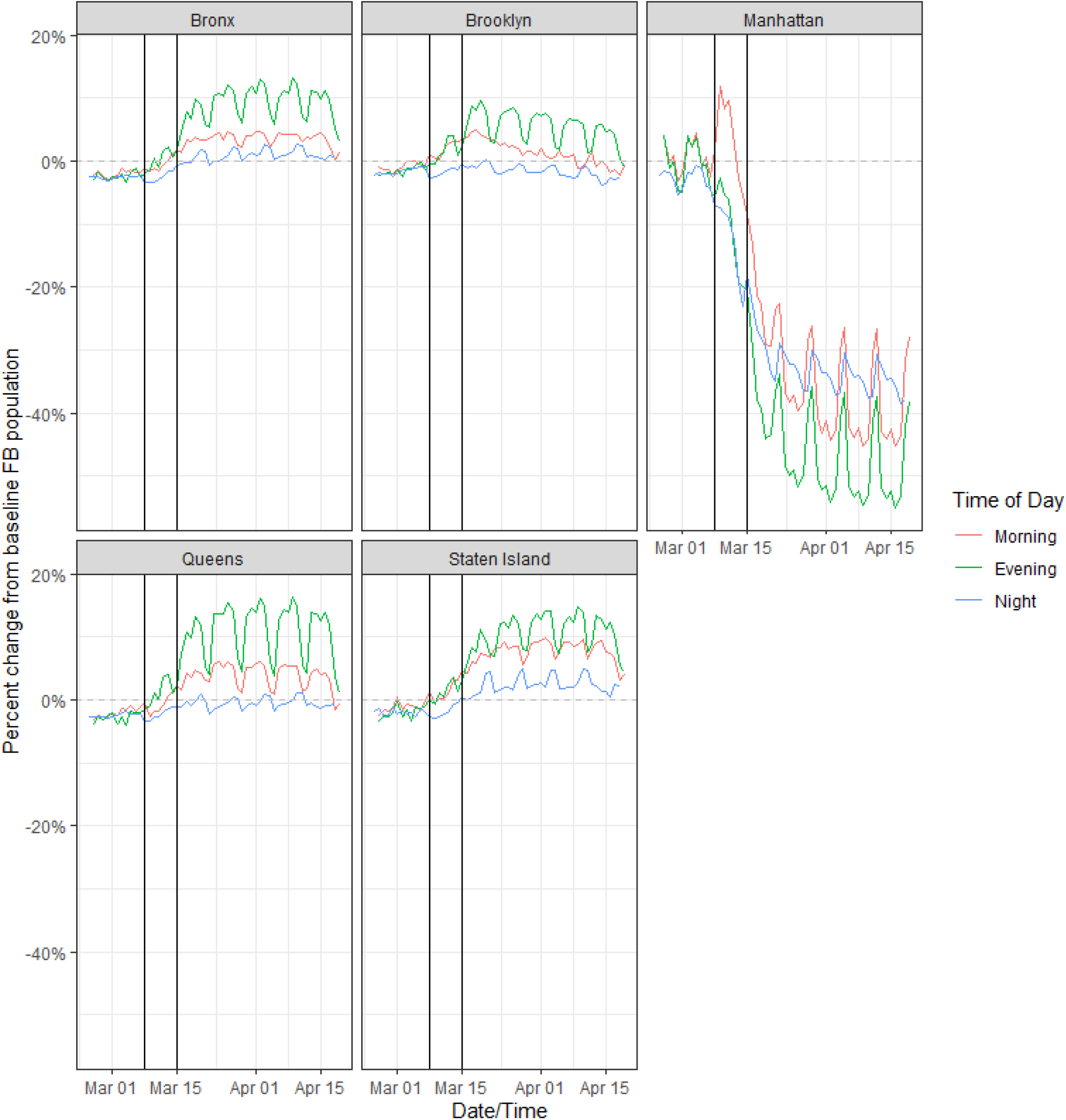
Percent change in population of Facebook users per region by day for boroughs in New York City, divided into times of day. The leftmost vertical black line is March 8th, 2020, the day that a number of schools began announcing closures. The rightmost vertical black line is the day that Governor Cuomo of New York ordered all New York City Schools closed.

The decrease in daytime population in Manhattan was not only driven by fewer workers coming into the city during the day, but also an exodus of residents out of the city overall, characterized by a decrease in nighttime population. In Figure 2 we evaluated this depletion of nighttime city populations, indicative of reduced residency, not just daytime activity, across other cities in the US for which data were available around the same time period, and we found a similar decrease in nighttime population. In New York City, the decline in residents overall was driven primarily by the exodus from Manhattan, and we expect similar heterogeneities may characterize population changes in other cities.

**Figure 2.**
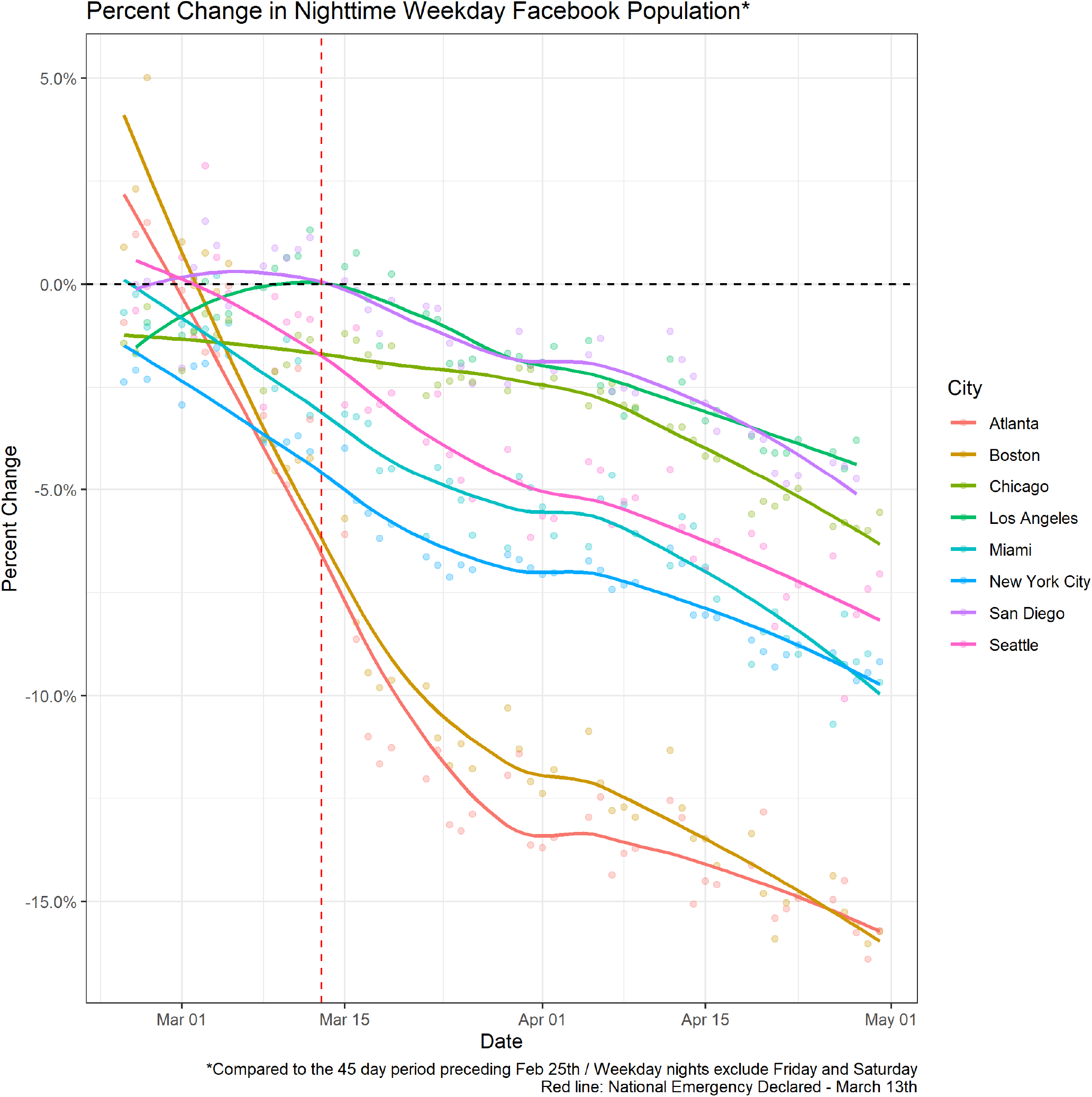
Percent change in weekday nighttime population of Facebook users by city. We can see that all cities included in the Facebook sample experience a decrease in nighttime population over the period of interest.

We further evaluated changes in urban versus rural populations on a national scale in France, Spain, India, and Bangladesh; four countries for which Facebook data pipelines were available to cover the timing of lockdowns. In Figure 3 regions in these countries are divided into five equally sized quantiles of nightlight 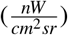 [10], which correspond to population density and reflect the urban-to-rural gradient. In each country, to varying degrees, there was a consistent decrease in population in areas with the highest nightlight intensity (urban centers) and a reciprocal increase in population in less electrified regions (more rural areas).

**Figure 3.**
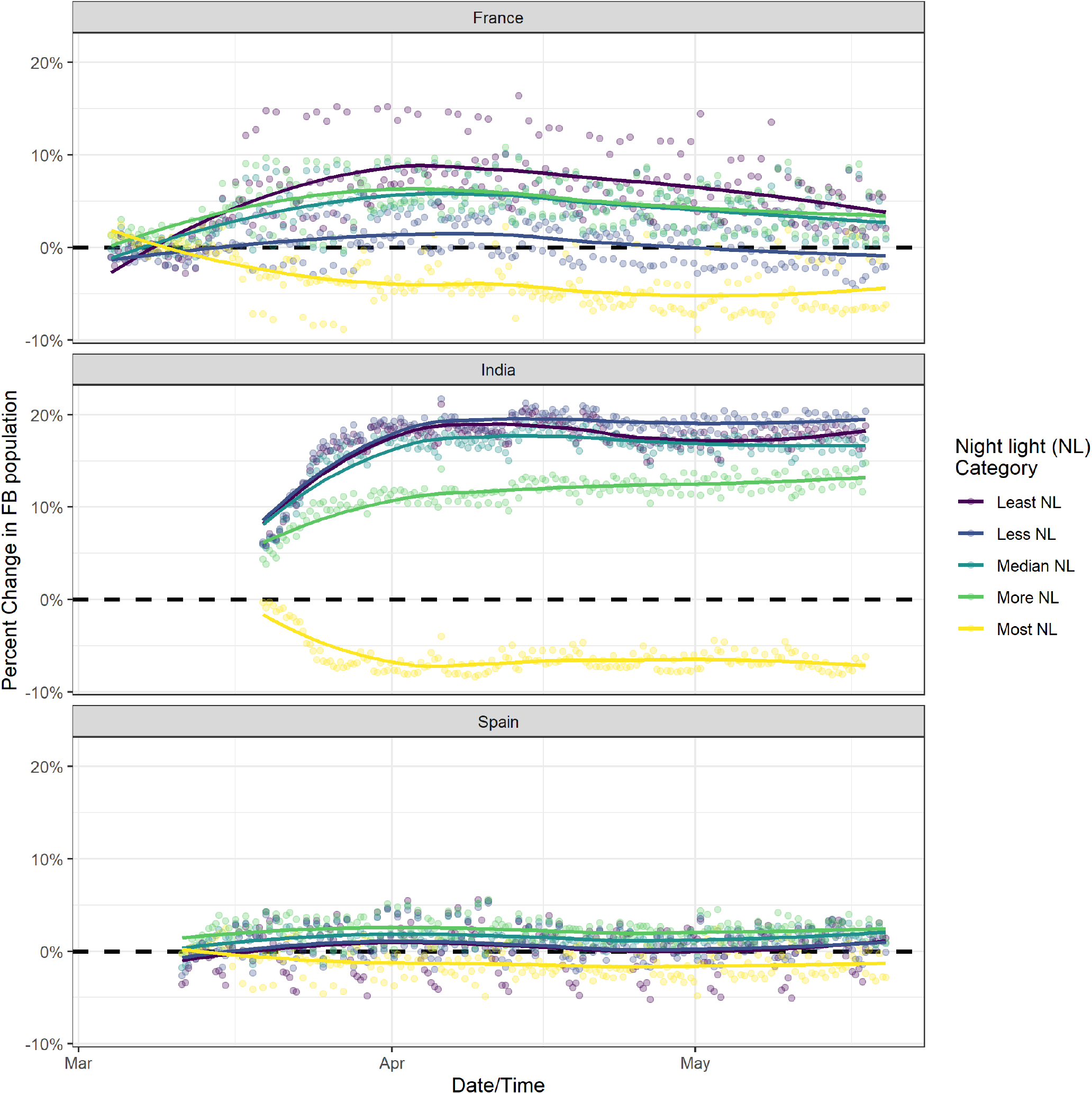
Percent change in population of Facebook users categorized by five equally sized quantiles of nightlight by country with data aggregated at the ADMIN3 level of spatial granularity.

In Bangladesh, we find a substantial decline in population in areas with the highest nightlight intensity - primarily in the capital, Dhaka, and areas with a high concentration of garment factories. The announcement of the lockdown in early March, and the closing of the garments industry, was followed by large movements of people from these densely populated urban areas to more rural areas [11, 12, 13]. Figure 4 shows the striking pattern of population decline in urban areas in March, followed by a gradual increase as the garments industry and other workplaces opened in late April. We are unable to use Bangladesh data for movement analysis or compare it directly to data from India, France and Spain as the spatial granularity of Facebook data provided is of poor resolution.

**Figure 4.**
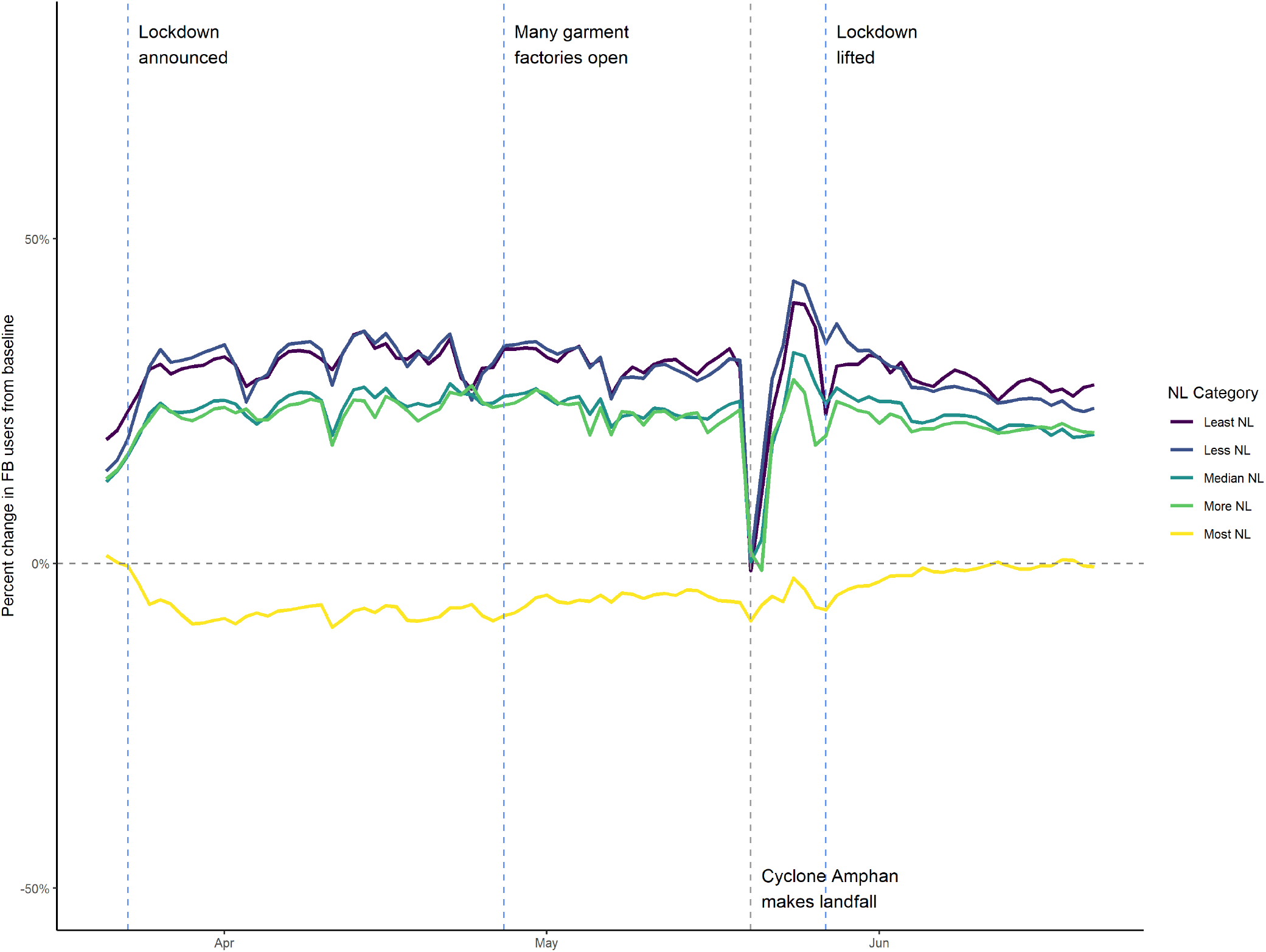
Percent change in population of Facebook users categorized by five equally sized quantiles of nightlight in Bangladesh with data aggregated at the ADMIN2 level of spatial granularity.

### Change in travel patterns

To better understand these changes, we evaluated the distribution of distances traveled around the country before, during, and after the lockdown (Figure 5). France announced a lockdown on March 16th and implemented it the next day while Spain implemented a state of alarm on March 13th and halted all non-essential activity on March 28th. In both countries, the vast majority of travel is local, but in France we see an immediate decrease in activity on the day that lockdown was implemented, while in Spain we see an initial decrease in long distance travel with a much more severe decrease once all non-essential travel was restricted. It is important to note that even with lockdown measures, the baseline rate of local travel remains generally unchanged, at times including trips of 50 kilometers or more in both France and Spain.

**Figure 5.**
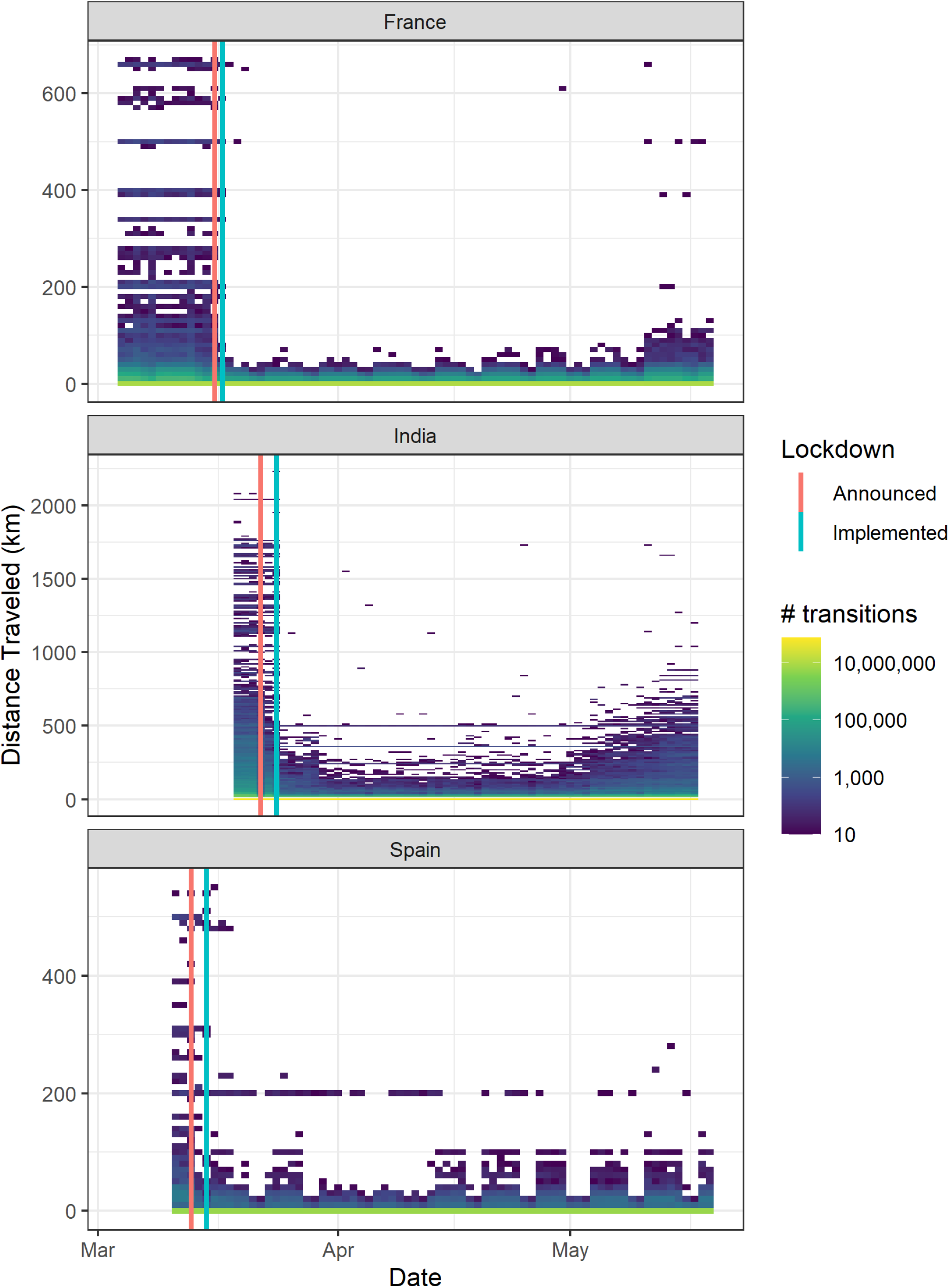
Number of trips made by distance. Here we can see that in all three countries, long distance trips decreased significantly in the immediate aftermath of lockdown implementation. However, local travel, including trips that ranged from 50-200 km remained during lockdown. This data is not available for Bangladesh.

### Pre-lockdown travel surges lead to faster and further initial spread of the epidemic

To examine the epidemiological implications of these behavioral responses to lockdowns, we implemented a metapopulation model reflecting the general behaviors we measured in the Facebook data. As shown in Figure 6, which depicts results from an epidemic without a lockdown or other interventions, we initiated the epidemic in an “urban” center (identified by a black outline) with a higher population density and evaluated the epidemic spread across all other “non-urban” areas, with travel determined by a gravity model of movement. We then varied travel and infection dynamics based on timing in relation to lockdown announcements and implementations (Figure 7). In the simulations, lockdowns affect behavior in two ways: first, between announcement and lockdown implementation contact rates within populations (*β*_1_) temporarily increase, and subsequently decrease once the lockdown takes effect (*β*_2_). Second, travel from urban to rural locations also changes prior to (*α*_1_) and following (*α*_2_) lockdown. We evaluated each possible parameter combination against a relative baseline where this is no travel surge and no increased contact rate during the period between lockdown announcement and implementation.

**Figure 6.**
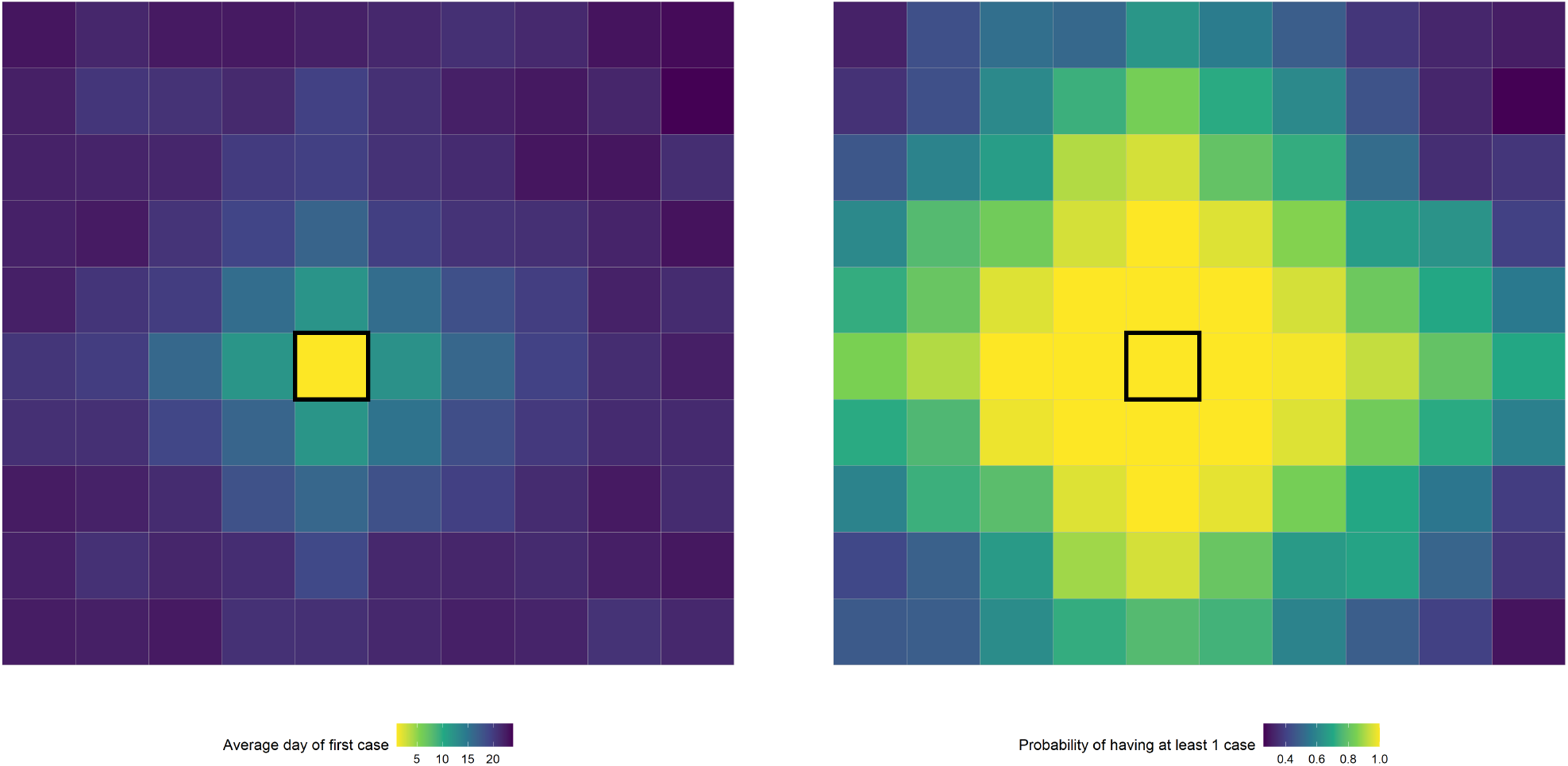
Results of a simulation evaluating: probability of at least one case by 30 days (right) and the average day that the first case appeared (left). The location outlined in black is the “urban” center with a larger population size and density. All other locations are “non-urban” with the same population size and density.

**Figure 7.**
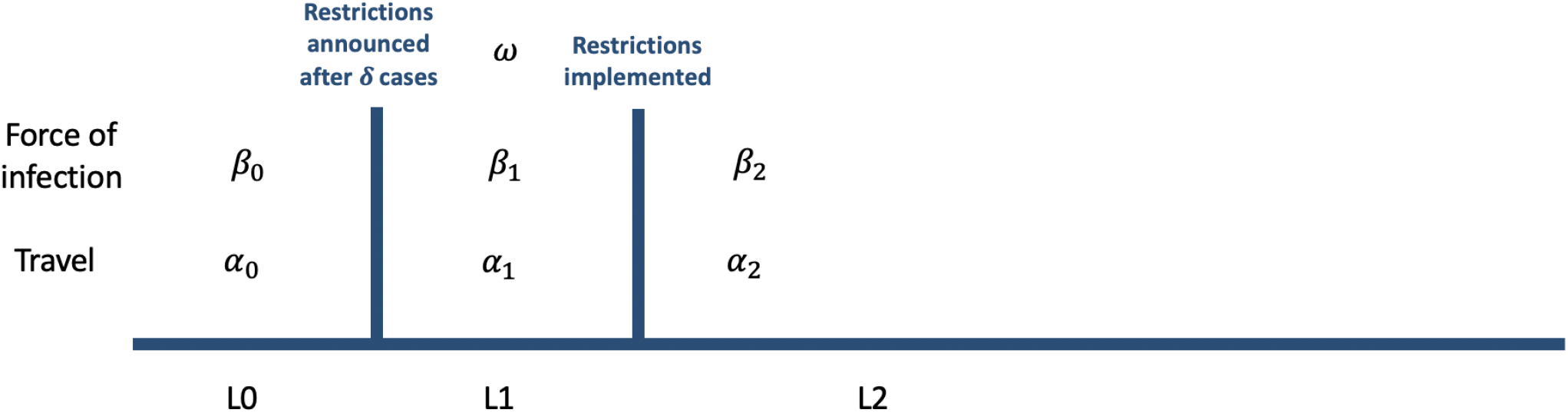
Diagram of the lockdown process and the values used for disease generation in communities and movement between communities.

While lockdowns decrease the spread of the epidemic if they are maintained effectively over time, travel surges at the beginning of an epidemic can increase exportation of cases out of the epicenter (Figure 8). In fact, travel surges can initially spread the disease faster than if no lockdown at all had been implemented. Changes in contact rates (*β*_1_) and travel (*α*_1_) following the announcement of a lockdown cause this increased rate of exportation of disease, with the former contributing more than the latter; however, there is a clear multiplicative effect as seen in Figure 9. Figure 9 (left) describes the relative probability of having an outbreak in a region within the first 30 days of the simulation, compared to a scenario where there is no change in *α* and *β* during the *L*_1_ period. This highlights the overall risk that communities face over the course of the epidemic, as well as the speed of outbreak spread. Given the novel nature of SARS-CoV-2 we have defined the detection of a single case in a given location as the clearest metric of this aspect of an epidemic, since stochastic localized outbreaks and return travel would subsequently complicate the spread of the virus. Figure 9 (right) evaluates the percent change in the number of days until an outbreak occurs, compared to the baseline scenario. This demonstrates the relative speed with which an epidemic is able to seed surrounding communities. As contact rates and travel increase, there is a corresponding increase in seeding of epidemics in new locations, as well as faster spread to all locations. This occurs because an increase in *β*_1_ results in a larger number of local cases available for travel while an increase in *α*_1_ results in an increased overall probability of those cases traveling.

**Figure 8.**
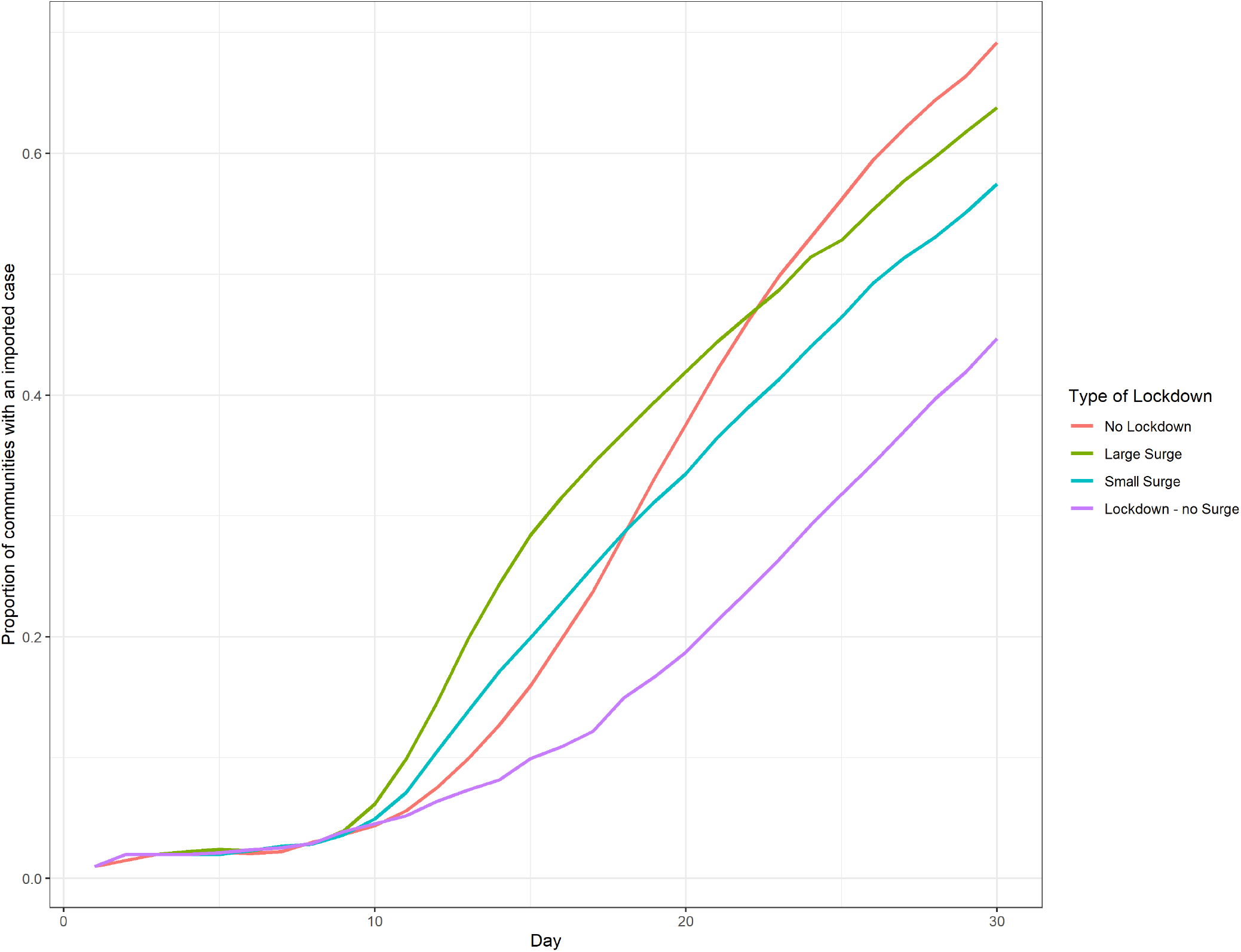
Average proportion of communities with an imported case. Initially the epidemic spread quicker in simulations with a large or small surge, however, simulations with no lockdown result in a larger overall epidemic size and eventually spread more rapidly.

**Figure 9.**
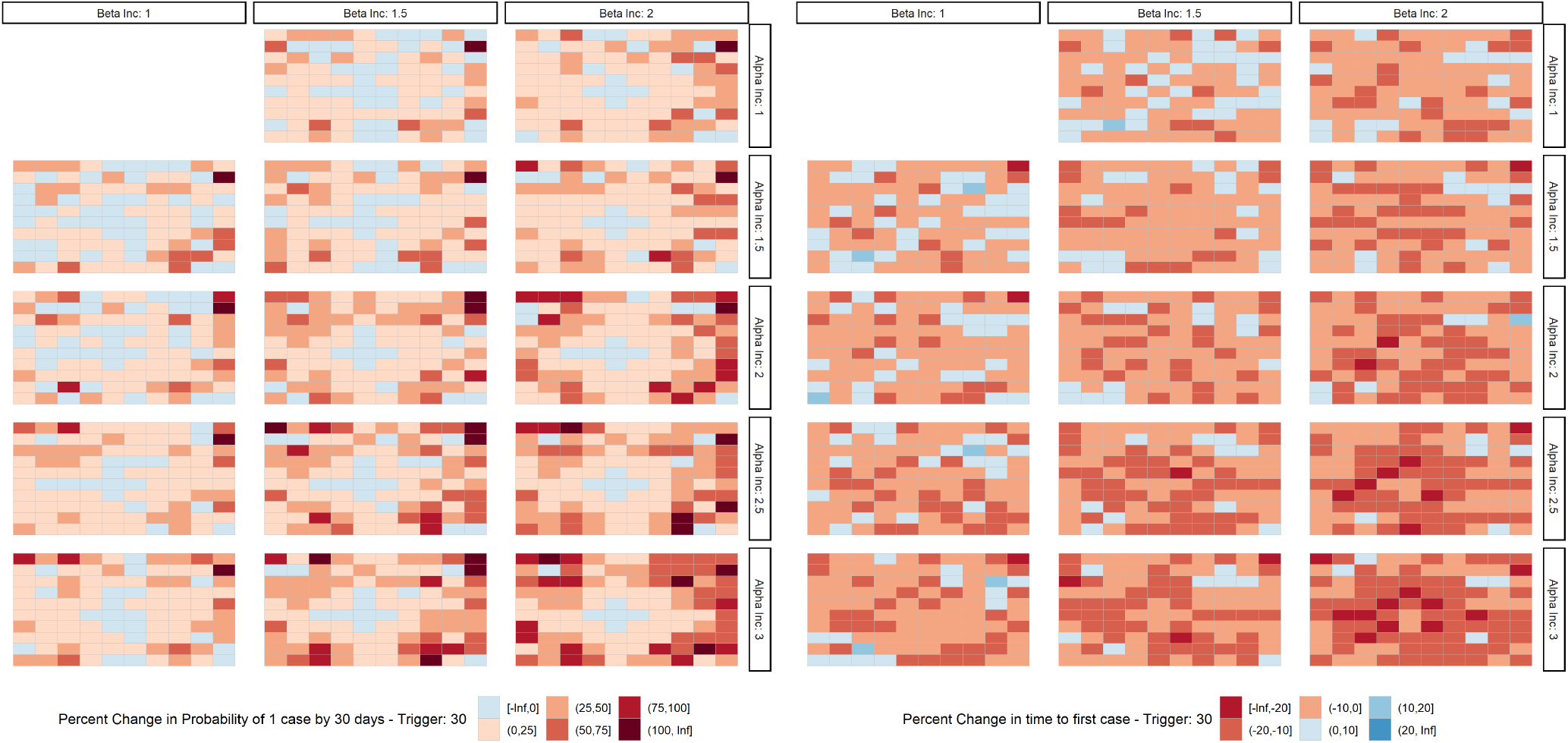
Percent change in probability of having at least 1 case by 30 days (left); Percent change in the number of days till the first case (right).

We evaluated the probability of travel (*α*_0_) under varying parameter values in the null model (i.e. no change in movement due to lockdown) with the goal of simulating a depopulation of the location that served as the urban center that was similar to what we found empirically in Figure 2. Across a variety of scenarios, an *α*_0_ of 0.01 (baseline daily travel probability) resulted in an at least 10% decrease in the population size of the urban center over the course of 60 days (Figures S1, S2 and S3).

### Rapid implementation of lockdowns after announcement decreases exported cases

Decreasing the time between announcement and lockdown implementation reduces the number of exported cases. As shown in Figure 10, an *L*_1_ period of 0 days resulted in no discernible increase in risk of an epidemic across all locations compared to the baseline. However, as we increased *L*_1_, the probability of having at least one case by thirty days increased in most non-urban locations. This effect was especially notable in rural locations far removed from the urban center. Importantly, the speed of the exportation of the epidemic was driven by both the duration of the *L*_1_ period and modification of the travel surge as defined by *α*_1_ and *β*_1_. With an *L*_1_ of 7 days, an *α*_*inc*_ of three and a *β*_*inc*_ of two, it is the locations that are closest to the urban center that have an exceptional decrease in the average number of days until the first case. The choice of timing between lockdown announcement and implementation must balance the increased risk of exportation from longer delays with the need to provide enough warning for people to adequately prepare for the lockdown.

**Figure 10.**
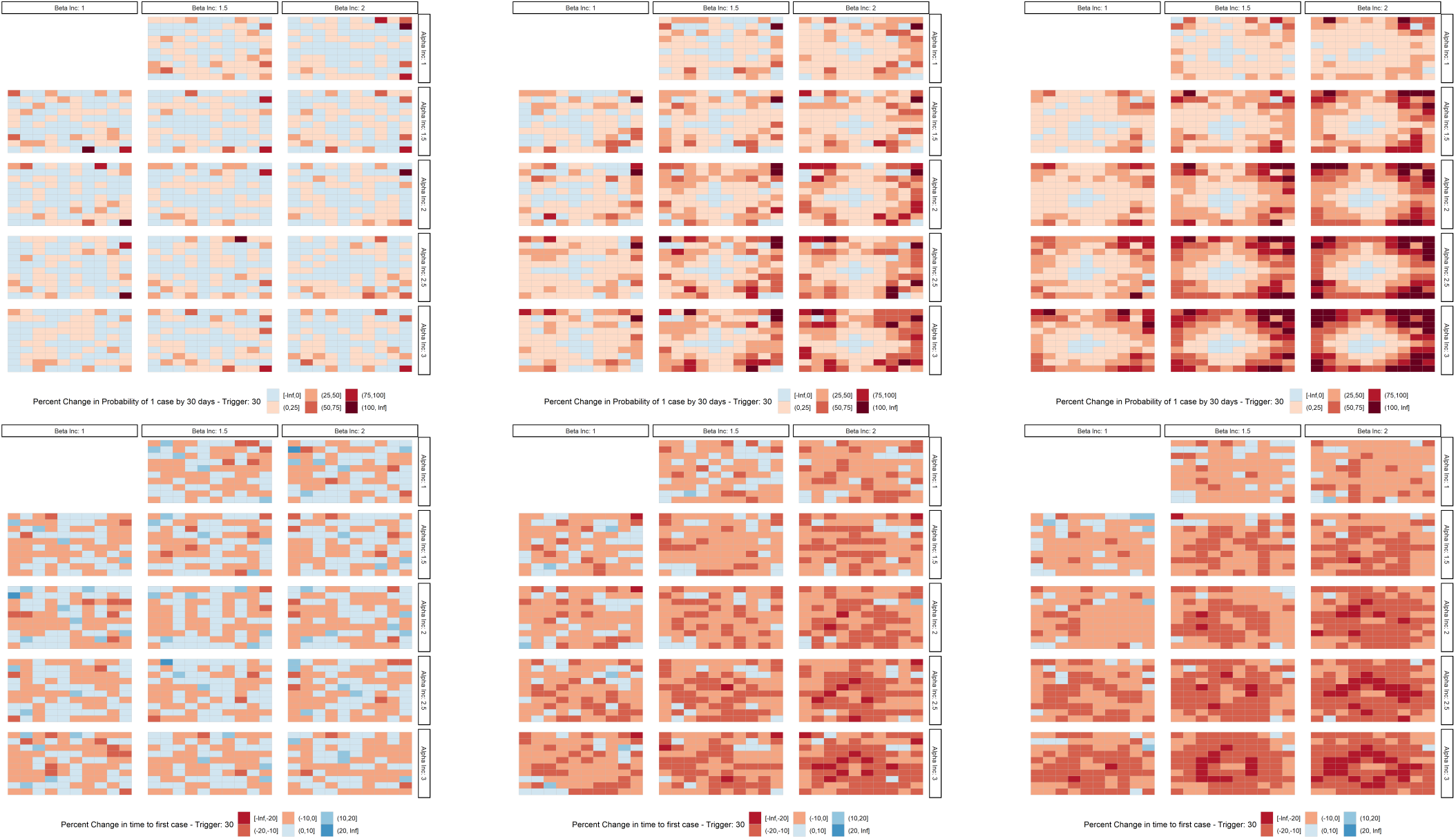
Top row, left to right: The percent change in the probability of having at least one case by 30 days in each location with an *L*_1_ period (time between lockdown announcement and implementation) of 0, 3 and 7 days respectively. Bottom row, left to right: The percent change in the average number of days till the first case in each location within an *L*_1_ period of 0, 3 and 7 days respectively.

## Discussion

### Exportation to rural areas is driven by travel and local contact rates

Human behavior in response to announcements of restrictions affects the trajectory of outbreaks. Both travel (*α*) and contact rates (*β*) play a key role in the increased risk of exportation of cases to non-urban locations following announcement of a lockdown. A temporary increase in local contact rates and mobility results in more epidemic seeding in rural areas compared to if the lockdown were implemented without these increases (Figure 8). Importantly, *α* drives the speed of the epidemic and greatly reduces the time until the first case in locations close to the urban center. The effect of *α* is modulated by the duration of the time between announcement and implementation (*L*_1_ period), with an increase in both probability of having an epidemic and the average time until the first case. The increase in risk is greater in regions that are further removed from the urban center as they have the lowest risk at baseline; however, the average time until the first case in these locations does not decrease nearly as much as those of locations closer to the urban center. This highlights the importance of proximity in our gravity model driven mobility network.

### Messaging surrounding the lockdown is key to reducing unintended consequences

This study shows that individually, the local contact rate, travel out of urban centers, or delay between announcement and implementation of a lockdown do not greatly affect the risk of case exportation and the propagation of the epidemic. However, the multiplicative effects of these factors can be dramatic. To decrease the probability of exportation, public health authorities must consider:

1. Messaging on how to prepare for lockdowns and expectations of the local supply chain to decrease instances of panic buying and hoarding, thereby decreasing the spike in local travel immediately preceding a lockdown [14].
2. Decreasing the window of time between announcement and implementation of lockdown policies to reduce both the local contact rate and the probability of out migration while balancing the needs of the population. Regardless of the duration of *L*_1_, lockdown policies which decrease *α* and *β* always resulted in smaller epidemics by the end of the study period (Figure 11).
3. Communicating the exportation risks associated with migration out of epidemic regions and coordinating between locations to reduce the risk of local transmission from an imported case [15]. This could include planning and resource allocation for increased surveillance and testing to account for the potential excess risk.
4. Providing the resources needed for people to stay. Strategies for mitigating travel surges will greatly depend on the reasons behind people’s movement. Movement of people back home from urban centers due to sudden lack of work from the pandemic [11] will require different interventions and messaging than people choosing to leave crowded cities for more remote second homes [16].

**Figure 11.**
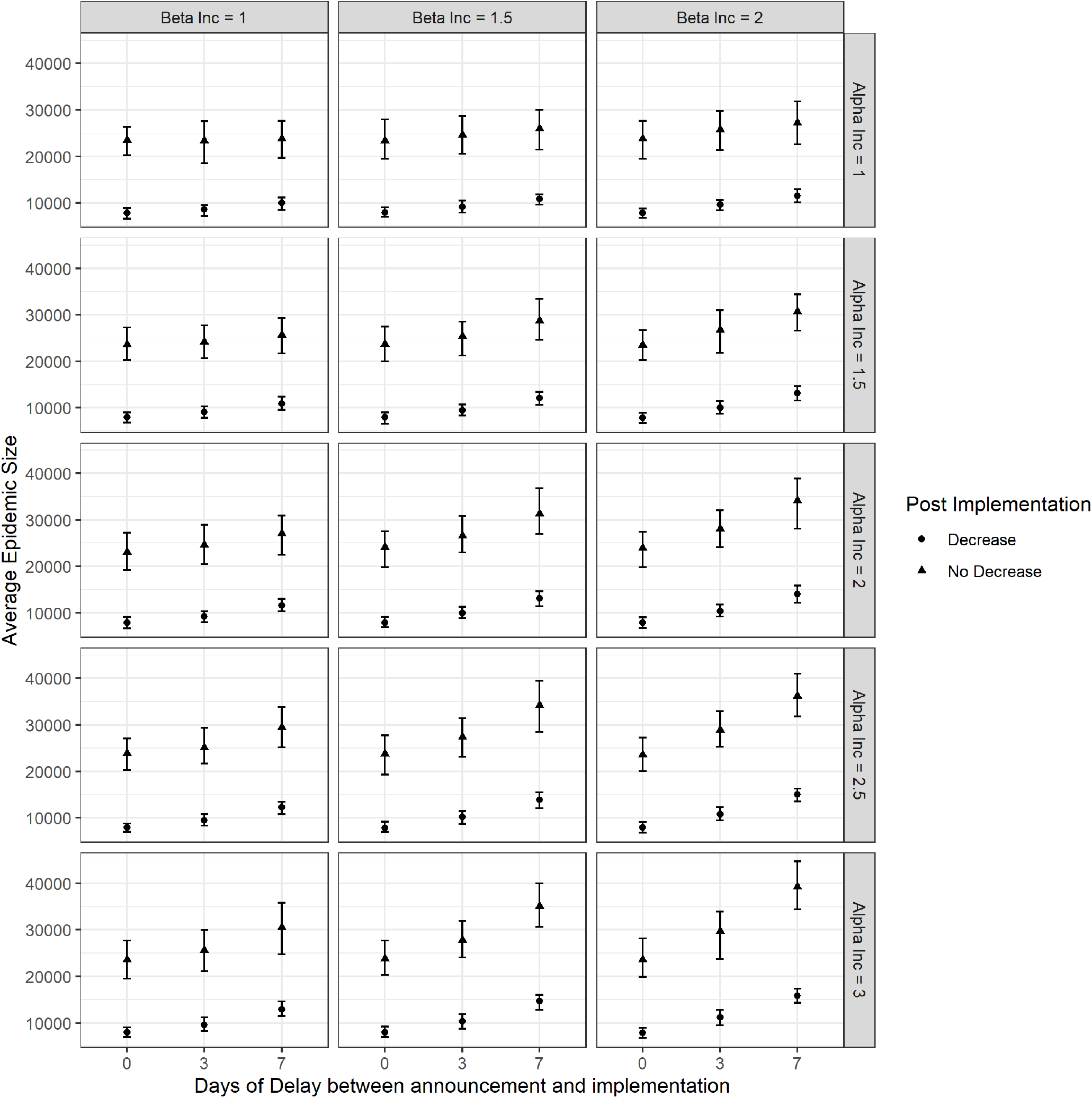
Overall epidemic size as over varying parameters. In all situations an epidemic with a lockdown (ie where there is a decrease in post lockdown travel) results in a smaller total epidemic size.

### Supporting epidemiologic data

Mobility and epidemiologic data support the findings from our model of increased exportation and seeding of epidemics as a result of travel surges. The mobility data provide evidence that lockdown announcements and implementation trigger travel surges and changes in travel patterns relative to pre-lockdown movement. In the simulations, we have shown how these changes in mobility can lead to increased introductions of cases in rural areas. Similarly, genomics analyses have found that many outbreaks across the United States were seeded by travelers from New York City [17]. Case data from Spain [18] also show increases in cases across a wide range of locations following the lockdown implementation. While many factors likely contributed to the similarities in the epidemic curves across locations, the increase in travel in the mobility data suggests seeding from urban areas may have played a role.

### Implications for Rural Areas

There are longstanding, global disparities in access to healthcare between urban and rural areas [19, 20], which have been further exacerbated by the SARS-CoV-2 pandemic [21]. If successful at reducing contact rates and travel, lockdowns can be an important tool for slowing the spread of an epidemic. However, the initial surges the lockdowns catalyze can cause the epidemic to spread further and faster to places outside of urban centers that may be less equipped to deal with an epidemic. Travel surges thus necessitate increased surveillance, testing, and treatment in areas that historically are understaffed and under resourced.

Through the course of a partially controlled epidemic in a susceptible population, our simulation shows that rural areas will still be affected, albeit at a later date (Figure 12). Appropriate messaging to decrease the spike in local contact rates and exodus out of epidemic areas along with inter-region coordination of movement can help decrease the burden of disease experienced by rural areas.

**Figure 12.**
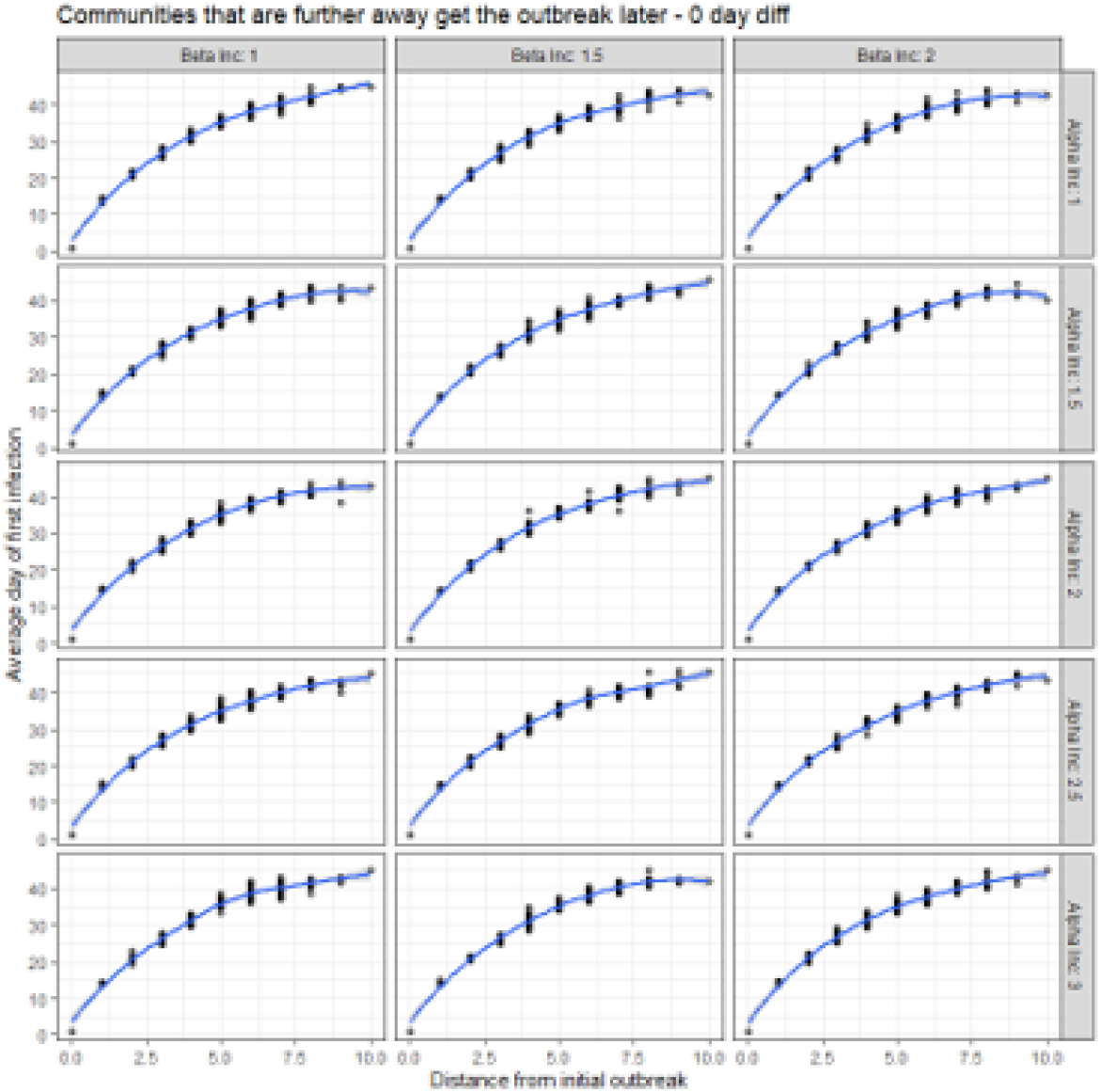
Average time of first case. Communities that are further away from the urban center are generally seeded later.

### Limitations

Many simplifying assumptions were made in the simulation model, including homogeneous mixing within locations on the lattice, a gravity model for connectivity, and the inclusion of only one urban center. Additionally, we assumed transmission dynamics were the same between symptomatic and asymptomatic individuals. Individuals in the I compartment are not able to travel immediately upon entering the I compartment, which may underestimate the amount of travel that would occur prior to symptom onset; however, given that those in A are able to travel, this likely will not impact the overall dynamics. We further assumed that increases in movement observed in the data following lockdown announcements coincided with increased contact rates, particularly in light of the anecdotal evidence of “panic buying”. However, in future outbreaks, interventions such as masks and social distancing, which were not consistently implemented in many places when lockdowns were first initiated, may reduce the correlation between movement and contact rates. Finally, the mobility analyses absorb the limitations of the Facebook data, which are limited to Facebook users with location services enabled. Despite these limitations, our results highlight the need for careful implementation of lockdowns to mitigate their potential unintended consequences.

## Methods

### Mobility Data

Facebook’s Data for Good team developed and provides access to the Geoinsights portal to provide movement and population level data in response to crises [22]. This interface allows researchers and response workers to request aggregated and anonymized datasets generated by an open cohort of individuals who are: 1) Facebook users; 2) have a smartphone, and; 3) are providing information through the Facebook app by having location services enabled. Data are requested for a geospatial region and defined by a spatial bounding box. For this analysis we used the movement and population datasets.

When the data aggregation pipeline is initiated, all individuals who are in the cohort described above and inside the bounding box contribute information to the datasets. For each user, location information is collected, and user location is categorized to Bing Tiles. The resolution of the Bing Tiles used varies by type of dataset with population data being offered at a higher resolution than movement data due to computational restrictions. Data are then aggregated into 8-hour bins. Population is determined by the modal location for each individual during this 8-hour bin. Movement for a given 8-hour bin is defined as a vector of transition with the destination being the modal location in the current 8-hour bin and the origin being the modal local for the preceding 8-hour bin. For each population tile and movement vector, Facebook provides a baseline which is calculated as the average number of users who were categorized as being in a given location (population) or who had made a given directional transition (movement) during the baseline period, conditional on day of week and time of day. The baseline period is defined as the 45-day period preceding the initiation of the pipeline for movement data and the 90-day period preceding the initiation of the pipeline for the population data.

### Selection of data sources

On February 27th, Facebook’s Data for Good team initiated the data collection pipeline for major cities in the United States of America. In the following weeks bounding boxes, and subsequent pipelines, were generated for regions as requested, including internationally. Our analyses are constrained to the locations with available data for the relevant time periods, and we use a combination of Facebook mobility data and nightlight data in different areas, as described below.

We restricted our sub-city analysis to New York City as 1) there are clear geographic borders (boroughs) with heterogeneity in the demographics of the population and land use in each region, 2) there were a large number of users included in the Facebook data set for each region, and; 3) the boroughs are of a large enough spatial scale to allow Facebook to capture highly granular movement and population data. City level analyses were restricted to the United States as Facebook initiated a city specific data collection pipeline for select cities on February 27th, well before the implementation of lockdown measures. Country level analyses were restricted to Spain, India, and France as all three countries quickly implemented strict lockdown measures, and Facebook initiated data collection pipelines for the whole country before these measures were put into place.

### Model Initialization

To assess the impact of different lockdown implementations and travel restrictions, we developed a simple metapopulation model, consisting of 100 communities, evenly spaced on a ten-by-ten lattice. One community in the center represents an “urban” area with a higher population size and population density than the other 99 “non-urban” locations. We make the simplifying assumption that all non-urban locations are homogeneous in terms of size and density and only differ in their distance from the urban. We seed an epidemic in the urban center with five initial cases. Within each community, the epidemic follows a density-dependent stochastic Susceptible-Exposed- Infectious(Asymptomatic)- Infectious(Symptomatic)- Recovered natural history. At each time step, all susceptible individuals have a chance of infection from the infectious individuals in their community, based on the parameter beta (i.e., force of infection). Asymptomatic and symptomatic cases are assumed to have the same beta, meaning the only difference between them in the model is whether or not they show symptoms. Detailed parameters of the outbreak are listed in Table 1. Individuals that are symptomatic (I) or asymptomatic (A) proceed through their disease history and approximately 10% of each compartment are removed into the recovered (R) compartment each time step for an average recovery period of 10 days [23].

**Table 1.** Simulation Parameters.

| Parameter | Value |
| --- | --- |
| Number of communities | 100 |
| Size of urban center | 4000 |
| Size of "non-urban" areas | 2500 |
| Area of urban center | 4 |
| Area of "non-urban" areas | 10 |
| Number of initial infections | 5 |
| Latent period | 5 days |
| Infectious period | 10 days |
| Proportion symptomatic | 0.5 |
| $\alpha_0$ (travel) | 0.01 |
| $\alpha_{inc}$ | 1, 1.5, 2, 2.5, 3 |
| $\alpha_{dec}$ | 0.5, 1 |
| $\beta$ (force of infection) | 0.0015 |
| $\beta_{inc}$ | 1, 1.5, 2 |
| $\beta_{dec}$ | 0.5, 1 |
| $\omega$ (days between announcement and lockdown) | 0, 3, 7 |
| $\delta$ (cases to trigger lockdown) | 10, 30 |
| Time steps | 60 days |

Following the time step specific movement through the disease generation process individuals in each community are given a chance to travel. This travel is driven by three factors: 1) the probability that an individual travels out of a given community, *α*_0_; 2) the probability that an individual from community i travels to community j, given that they will travel out of community i, *p*_*i j*_ | *α*_0_ and; 3) the disease status of the individual. All individuals that are in the S, E, A and R compartments are able to travel. Here we assume that individuals who are symptomatic and infectious will self-isolate and not travel. We first calculate the number of individuals that leave each compartment in each community, and then distribute them into the same compartment in another community, depending on the probabilities described above. As seen in Figure 2 and 3, we see wide ranging levels of depopulation in urban areas. In the most acute cases, such as in Manhattan in Figure 1, we see an approximate 40% decrease in the nighttime population. However, in country level analyses this can vary significantly. We have tuned the *α*_0_ parameter in our model to result in an approximately 10% reduction in our “urban” population over the length of our model run. The value 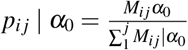 where *M*_*i j*_ is the i specific normalized value of a simple gravity model defined as:

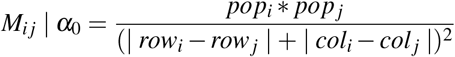

Here the values for row and col return the row and column number of the community in our ten-by-ten lattice. Given that an individual moves, the location that they move to is determined by a gravity model with locations that are closer and locations which are more heavily populated (i.e. the urban center) receiving a higher probability of travel.

### Timing and tuned parameters

We designed our model to describe three distinct periods of time: 1) *L*_0_, the period before any lockdown measures are announced or implemented, 2) *L*_1_, the period of time after announcement of lockdown, but before implementation; and, 3) *L*_2_, the period of time after the implementation of the lockdown (Figure 7). As described above, the initial parameters of the disease generation process and movement were controlled with *α*_0_ and *β*_0_, which were tuned empirically. We varied six parameters which influenced these initial parameters to evaluate the impact of differential implementation of lockdowns as shows in Figure 7.

- *α*_*inc*_: A multiplicative factor which describes the increase in *α*_0_ during the *L*_1_ period resulting in *α*_1_. We used this variable to simulate the increase in movement out of urban areas. *α*_*inc*_ is assumed to be constant throughout the *L*_1_ period.
- *α*_*dec*_: A multiplicative factor which describes the decrease in *α*_0_ during the *L*_2_ period resulting in *α*_2_. We used this variable to simulate the reduction in movement between all locations resulting from the implementation of a lockdown.
- *β*_*inc*_: A multiplicative factor which describes the increase in *β*_0_ during the *L*_1_ period resulting in *β*_1_. We used this variable to increase the force of infection in areas where an epidemic had already started to simulate the increase in the contact rate between individuals due to greater local movement.
- *β*_*dec*_: A multiplicative factor which describes the decrease in *β*_0_ during the *L*_2_ period resulting in *β*_2_. We used this variable to decrease the force of infection in the areas where an epidemic had already started to simulate the decrease in the contact rate likely after the implementation of lockdown measures.
- *δ* : The number of local symptomatic cases necessary for announcement and implementation of lockdown measures. Here we assumed that all symptomatic cases were immediately identified.
- *ω*: The amount of time between announcement of a lockdown and implementation.

### Metrics

We simulated the stochastic epidemic 100 times. In each of the 100 communities, we calculated the proportion of simulations in which that community had at least one case by day 30. We also calculated the average time to first infection across simulations in each community. We compared these two metrics across variations of the six parameters described above. For our primary analysis we held *δ* constant as it did not directly affect our question of interest. We subsequently varied *δ* to evaluate the sensitivity of our model.

## Data Availability

Code is available on github. Due to the Data Sharing Agreement with Facebook, readers can not access the original Facebook data used in this study.

https://github.com/nish-kishore/COVID-travel

## Supplementary Materials

**Figure S1.**
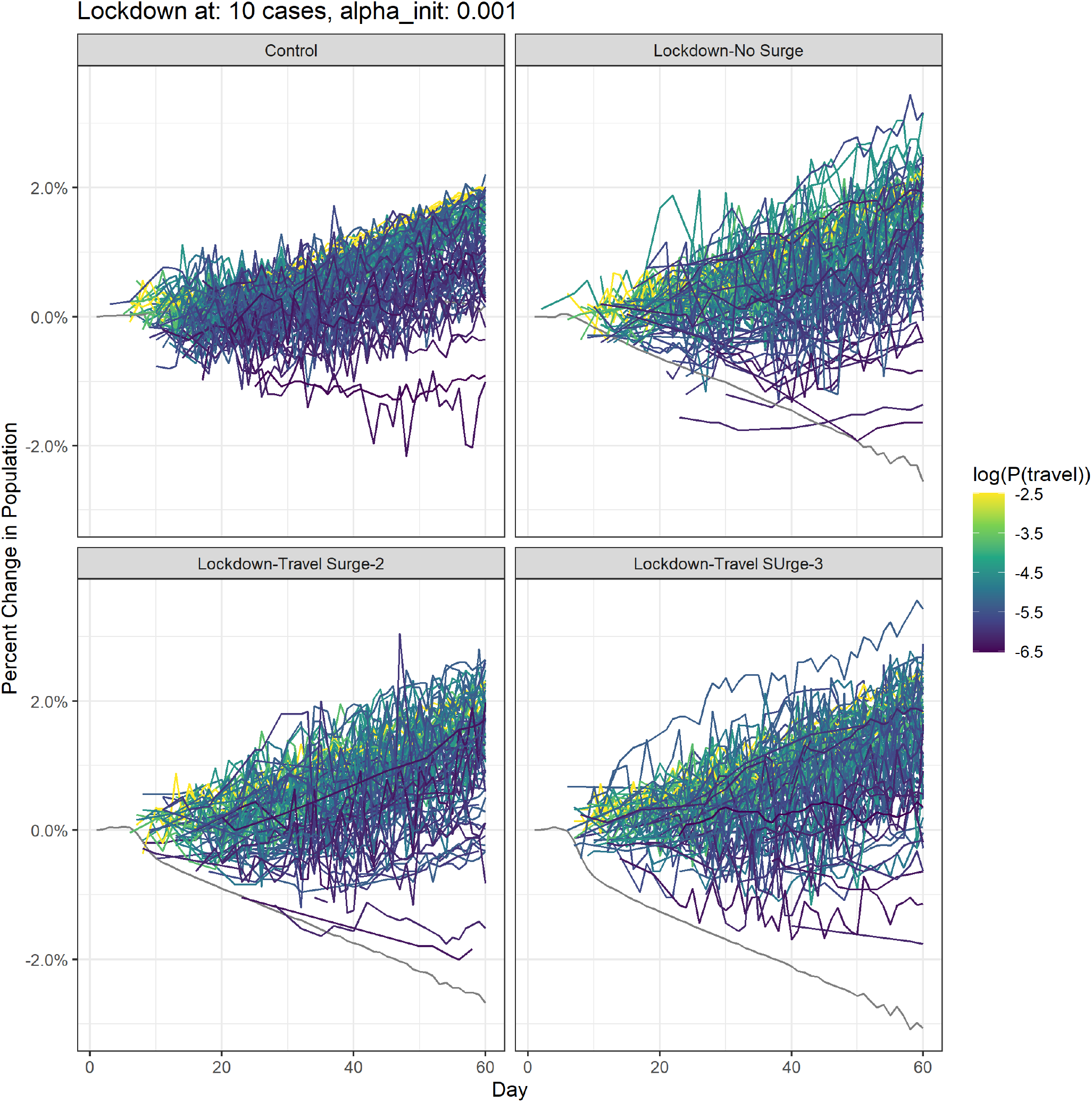
Depopulation of urban center with *α*_0_ of 0.001.

**Figure S2.**
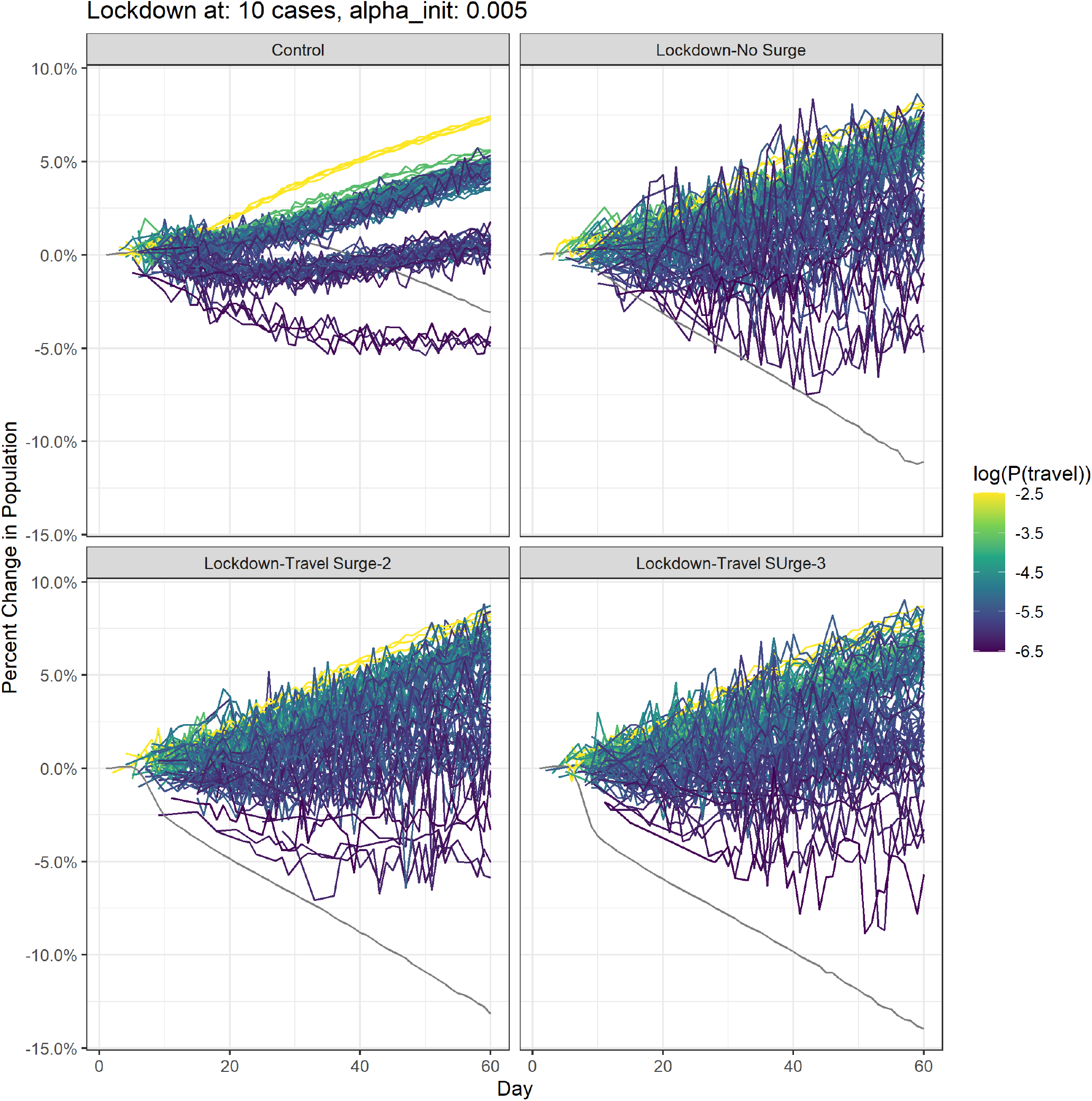
Depopulation of urban center with *α*_0_ of 0.005.

**Figure S3.**
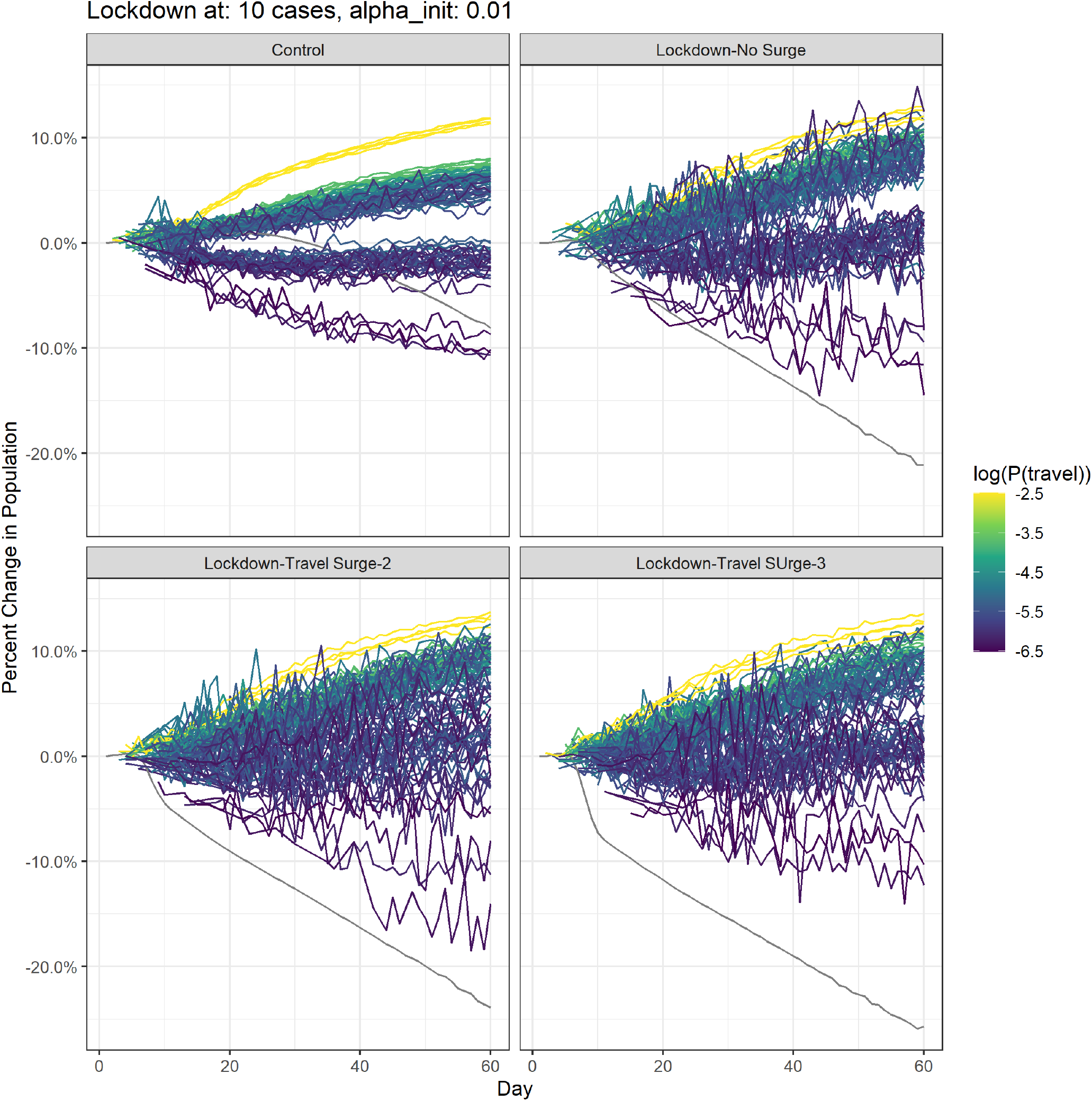
Depopulation of urban center with *α*_0_ of 0.01.

